# Residential clustering of COVID-19 cases and efficiency of building-wide compulsory testing notices as a transmission control measure in Hong Kong

**DOI:** 10.1101/2022.10.12.22280904

**Authors:** Benjamin R. Young, Bingyi Yang, Peng Wu, Dillon C. Adam, Jessica Y. Wong, Faith Ho, Huizhi Gao, Eric H. Y. Lau, Gabriel M. Leung, Benjamin J. Cowling

## Abstract

**Background:** Despite relatively few reports of residential case clusters of COVID-19, building-wide compulsory testing notices on residential apartment blocks are frequently applied in Hong Kong with the aim of identifying cases and reducing transmission.

**Methods:** We aimed to describe the frequency of residential case clusters and the efficiency of compulsory testing notices in identifying cases. The residences of locally infected COVID-19 cases in Hong Kong were grouped to quantify the number of cases per residence.

Buildings targeted in compulsory testing notices were matched with the residence of cases to estimate the number of cases identified.

**Results:** We found that most of the residential buildings (4246/7688, 55.2%) with a confirmed COVID-19 case had only one reported case. In the fourth and the fifth epidemic wave in Hong Kong, we estimated that compulsory testing notices detected 29 cases (95% confidence interval: 26, 32) and 46 cases (44, 48) from every 100 buildings tested (each with hundreds of residents), respectively. Approximately 13% of the daily reported cases were identified through compulsory testing notices.

**Conclusions:** Compulsory testing notices can be an essential method when attempting to maintain local elimination (‘zero covid’) and most impactful early in an epidemic when the benefit remains of stemming a new wave. Compulsory testing therefore appears to be a relatively inefficient control measure in response to sustained community transmission in the community.

## INTRODUCTION

Hong Kong as other large cities in the world has a high population density and many residents live in high-rise apartment buildings where the median gross area is 430 square feet (40 square meters), with an average of 2.8 residents per household (1). The severe acute respiratory syndrome coronavirus 2 (SARS-CoV-2) has been hypothesized to transmit on some occasions within high-rise apartments vertically through the sewage stack (2), horizontally along corridors (3) as well as via outdoor routes through open windows and vents (4). While more than a third of documented transmission events occurred within households during the first nine months of the pandemic (5), there have been few large outbreaks involving multiple households in residential estates (2). This is in contrast to the 2003 outbreak of Severe Acute Respiratory Syndrome (SARS) when more than 300 of 1755 cases in Hong Kong occurred in 200 households in a large outbreak in six blocks of the Amoy Gardens housing estate, with infection potentially spread through the air via sewage drainage pipes (4, 6).

Hong Kong has experienced five epidemic waves over the course of the pandemic to date. The first four waves were caused by the ancestral SARS-CoV-2 strain and were controlled through a combination of public health and social measures (7), leading to 13,000 confirmed cases documented, and less than 1% of the population infected as indicated in a serologic study (8). However, a large fifth wave occurred in early 2022 with Omicron BA.2 and more recently Omicron BA.5 and BA.4, causing more than a million confirmed cases and more than 9000 deaths (9). While stringent public health and social measures have been implemented in the fifth wave, the suppressed epidemic wave never dies out. One control measure implemented in Hong Kong since November 2020 is the issuance of compulsory testing notices (CTNs) on residential buildings when one or more cases are identified among the residents of that building or when SARS-CoV-2 virus is identified in sewage samples collected from that building. When a CTN is issued on a particular building, all residents of that building are required to provide respiratory specimens (or stool specimens for children <3y) at community testing centres on specified dates (wave 4: within three days, wave 5: within two days and repeated on multiple dates). Starting in early 2021, a more stringent approach was used when a higher risk of residential transmission was suspected, some buildings would be issued a CTN and placed under a ‘restriction-testing declaration’ (RTD) as well, locking the building down for a period of time (usually for around 12 hours overnight) to allow all residents to be sampled for RT-qPCR test immediately, with residents not permitted to leave the building until all test results are obtained. The longest RTD occurred in a building for an 8-day lockdown in early 2022 at the start of the fifth wave (10). On a small number of occasions, all residents of a building with confirmed cases were required to undergo weeks of quarantine at designated facilities (11). The objective of this study is to investigate the extent of residential case clustering throughout the pandemic and the efficiency of building-wide CTNs and RTDs in the control of COVID-19 in Hong Kong.

## 1. MATERIAL and METHODS

### 1.1 Study period and population

We analyzed information on CTNs, RTDs, and individuals with laboratory-confirmed SARS-CoV-2 infection who lived in residential buildings, excluding institutions such as residential care homes. We excluded laboratory-confirmed cases that were classified as having been infected outside Hong Kong (“imported cases”). Our study period covered all local epidemics that occurred from early 2020 through to 15 February 2022. After the latter date, the rapid increases in number of daily cases precluded detailed information being reported on every case. We analyzed data on CTNs and RTDs after the implementation of these initiation of these policies i.e., after November 2020 for CTNs and after January 2021 for RTDs.

### 1.2 Data collection

Information on each eligible case was collected from a line list of COVID-19 cases including the report date, date of symptom onset, residential building address, virus variant causing infection and any known associated clusters. To analyse the variants of SARS-CoV-2, cases were grouped into four categories, the ancestral strain, Delta or Omicron variants of concern, or other/undetermined variants, dependent on the report date and any viral mutations sequenced. Cases identified in the first four waves were considered infections with the ancestral strain (12), infections in the fifth wave with a N501Y gene mutation in the virus RNA were determined to be the Delta variant and those with N501Y and T478K gene mutations were the Omicron variant. Cases identified after 6 February 2022 without sequencing data were assumed to be infected by the Omicron variant as it was the predominant variant by this time (7).

### 1.3 Cases and clusters per building

For each case, the residential address details were extracted and building addresses geocoded with the respective building geocoordinates. The number of cases and clusters per residential building were estimated using an algorithm developed to group the cases by unique residential building based on a combination of geocoordinates and address details (see appendix, residential grouping algorithm). We defined a case cluster as two or more epidemiologically linked cases.

### 1.4 Compulsory testing notices

From detailed CHP press releases we identified and geocoded the residential buildings targeted by CTNs during the study periods. For each CTN, we collected the justification and the initiation and closure dates of CTNs including the date(s) of mandatory testing. With regards to RTDs specifically, data on the total number of residents that underwent a COVID-19 test and the number of cases identified within the RTD were also collected.

To determine the number of reported cases associated with CTNs, the reported cases were matched with the targeted buildings using coordinates, address details, CTN issuance and mandatory test dates and case report dates. Cases matched with buildings in RTDs were limited to those reported on the day the lockdown was lifted as lockdowns would not be lifted until the COVID-19 test results were available for all residents. If the lockdown was extended for multiple days, cases reported during and up to one day after the lockdown were considered associated with the RTD. Cases identified from buildings receiving CTNs due to ‘confirmed case(s) in the building’ or ‘positive sewage samples’ were those reported one day after the initiation of CTN up to one day following the last date of mandatory testing. If a building had multiple CTNs open at once, the proximity of the case report date to the mandatory testing date would determine which CTN accounted for the case. If the mandatory tests shared the same date, the earlier CTN issued accounted for any cases identified during that period.

Cases matched with a CTN were further classified to distinguish those which could have been identified via contact-tracing alone versus cases that were determined likely to have only been identified via the CTN and therefore we considered “untraceable” (see appendix, ‘untraceable’ cases algorithm). The proportion of asymptomatic CTN-associated cases were also determined using the reported symptom onset data.

### 1.5 Statistical analysis

The daily number of CTN-associated cases aggregated on the mandated test date was divided by the daily number of local cases on the same date to calculate the proportion daily cases associated with CTNs, and the average daily proportion was weighted by the daily local cases. The RTD-test positivity per 100 residents tested was calculated by dividing the reported number of PCR-confirmed cases detected in RTDs by the reported number of residents tested and multiplying 100. To determine the daily RTD test-positivity, cases associated with a multi-day RTD were distributed amongst the days over the RTD following a normal distribution with a standard deviation of two, while the number of residents tested in the RTD was distributed evenly over the testing period. The ratios of CTN-associated cases and untraceable cases per 100 buildings targeted in CTNs and per 100 building-wide mandated tests were calculated and stratified by epidemic wave. The 95% confidence intervals for the ratios were calculated using the Clopper-Pearson Exact method. The cumulative proportion of CTN-associated cases coded as ‘untraceable’ was calculated by the sum of ‘untraceable’ cases divided by the CTN-associated cases within the period of interest. An ordinal logistic regression model was fit to determine whether there was a difference in odds of a higher number of cases (6-20, >20 cases) or clusters (2-5, >5 clusters) per building associated with the infecting variant compared with a lower number of cases (1-5 cases) and clusters (<2 clusters) per building.

As a sensitivity analysis, the residential grouping algorithm was checked by randomly sampling 100 buildings (50 from waves one to four and 50 from wave five) with reported cases and manually assessing all cases associated with each respective building. The sensitivity and specificity of the reported case and CTN matching algorithm were assessed by randomly sampled 100 matched and 50 non-matched cases, to manually check whether they were true matches or true non-matches, respectively, based on our pre-defined criteria.

Pearson-correlation coefficients were used to assess the relationship between the daily number of local cases and the SARS-CoV-2 test-positivity in RTDs. Similar methods were used to examine the relationship between the daily number of local cases and the number of buildings targeted in CTNs.

## 2. RESULTS

As of 15 February 2022, there had been 26,670 laboratory-confirmed COVID-19 cases in Hong Kong, which included 22,782 (85.4%) locally infected and 3888 (14.6%) imported cases. Among the local infections 9,382 occurred between Hong Kong’s first (22 January 2020) and fourth wave (up to 25 April 2021), while 13,323 occurred during the rising phase of the fifth wave (24 December 2021 – 15 February 2022) (Figure 1A).

**Figure 1.**
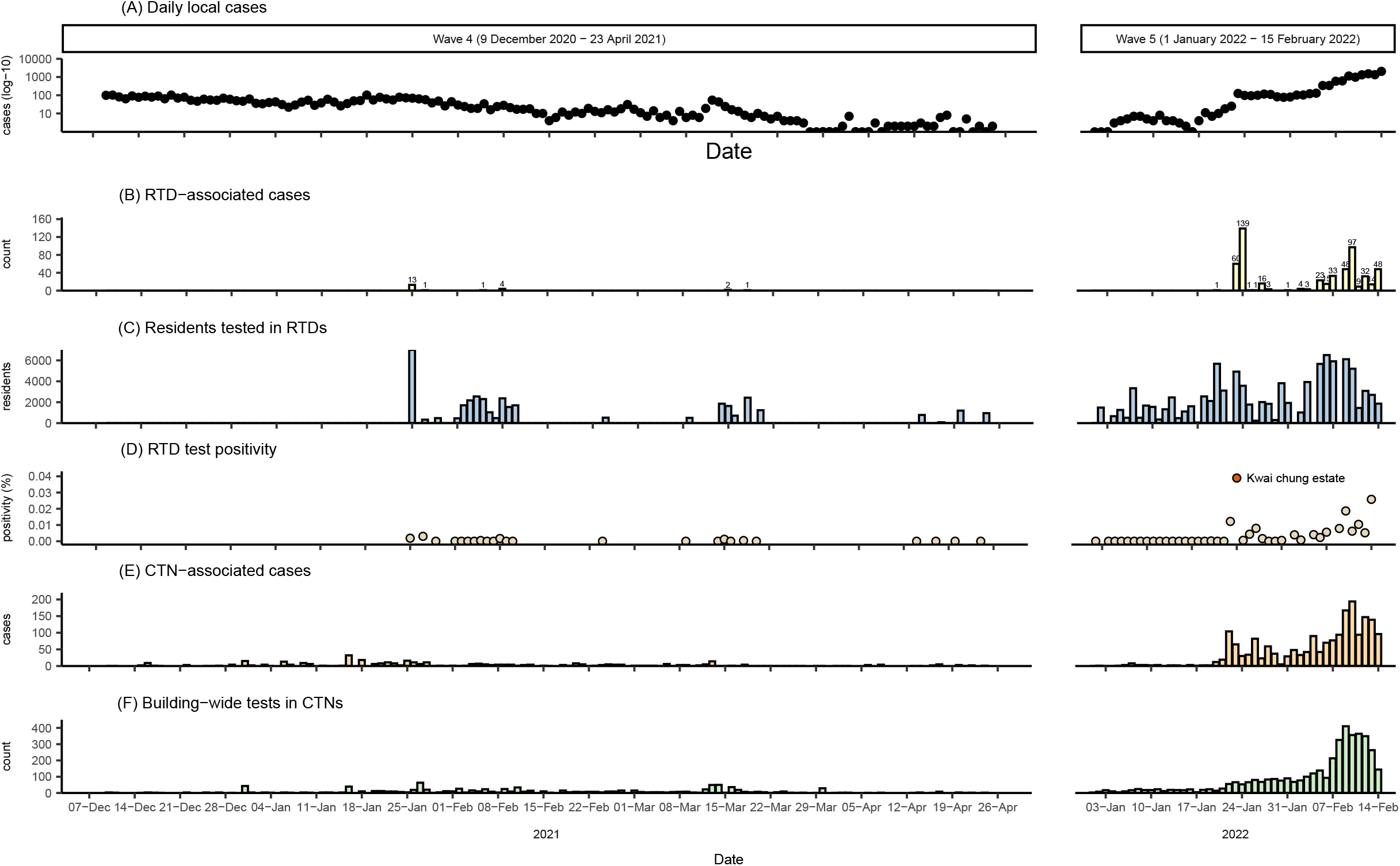
Panel A: the daily local cases (logarithmic scale) across the end of the fourth wave (9 December 2020 – 23 April 2021) and the rising phase of the fifth wave (1 January 2022 – 15 February 2022), during the period that the restriction testing declarations were mandated. Panel B: The daily cases associated with restriction testing declarations. Panel C: The daily number of individuals tested within each lockdown. Panel D: the daily test-positivity is shown, representing the number of cases identified out of the residents tested in RTDs. Panel E and F show the cases associated with building-wide compulsory testing notices and the daily number of buildings with a mandated test, respectively.

The first RTD was issued in January 2021 during the fourth wave. Forty RTDs occurred during the fourth wave targeted 305 residential buildings and 36,006 residents, with a mean test positivity of 0.04% (Figure 1B-D). The largest RTD in the fourth wave was on 23 January and lasted 40 hours (median duration of lockdowns: 14 hours) with over 3,000 staff members involved to test around 7,000 individuals. Within the first six weeks of the fifth wave, ninety-two RTDs were issued on 171 residential buildings and 98,603 individuals, resulting in 561 cases and a mean test positivity of 0.35% (variance = 0.82%) (Figure 1B-D). Between 19 January 2022 and 29 January 2022, six RTDs were issued across seven different buildings at the Kwai Chung Estate, with extended lockdowns and repeated testing. Over the ten-day period around 35,000 COVID-19 PCR tests were administered, impacting an estimated 12,000 residents, which were linked with at least 216 reported cases.

Figure 2A shows the relationship between the daily RTD test positivity rate (cases detected per 100 residents tested) and the daily number of local cases. The statistically significant (p-value <0.001) correlation coefficient moves closer to 1, from 0.59 to 0.76, if the large residential outbreak at the Kwai Chung Estate is removed (Figure 2A). However, cases linked to RTDs comprised just 4.0% (95% CI: 1.2%, 6.8%, weighted by daily local cases) of all cases confirmed each day and there was no statistically significant correlation between the two metrics (p=0.757) (Figure 2C).

**Figure 2.**
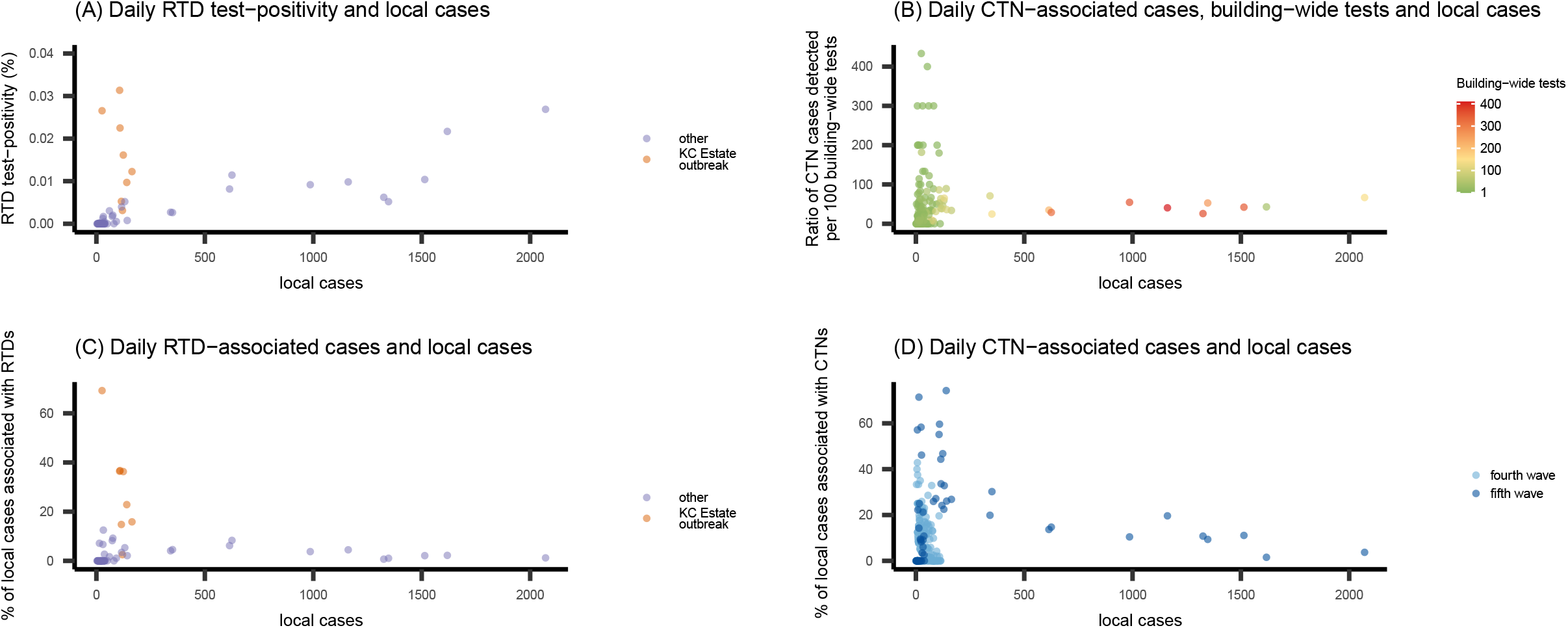
(A) The positive association between the daily test-positivity during restriction testing declarations (RTDs) (lockdown-associated cases/residents tested per lockdown) and the daily number of local COVID-19 cases identified in Hong Kong. (B) The relationship between the daily detection of compulsory testing notice (CTN)-associated cases per building-wide mandated tests, daily local cases, and building-wide mandated tests performed under CTNs. (C) The relationship between the percentage of local cases associated with an RTD and the daily local cases, and (D) The relationship between the daily percentage of local cases associated with a CTN and the daily local cases.

During the fourth wave 1057 CTNs were issued to 1321 residential buildings, and 345 cases associated with the CTNs were identified over a four-month period. In the first six weeks of the fifth wave, 4174 building-wide tests were mandated via CTNs across 1097 residential buildings and led to 1888 associated reported cases (Figure 1E-F). The justifications for issuing CTNs are displayed in Table 1, and CTNs triggered by other than the pre-determined reasons were excluded from our analyses (Table S1 & S2).

**Table 1.**
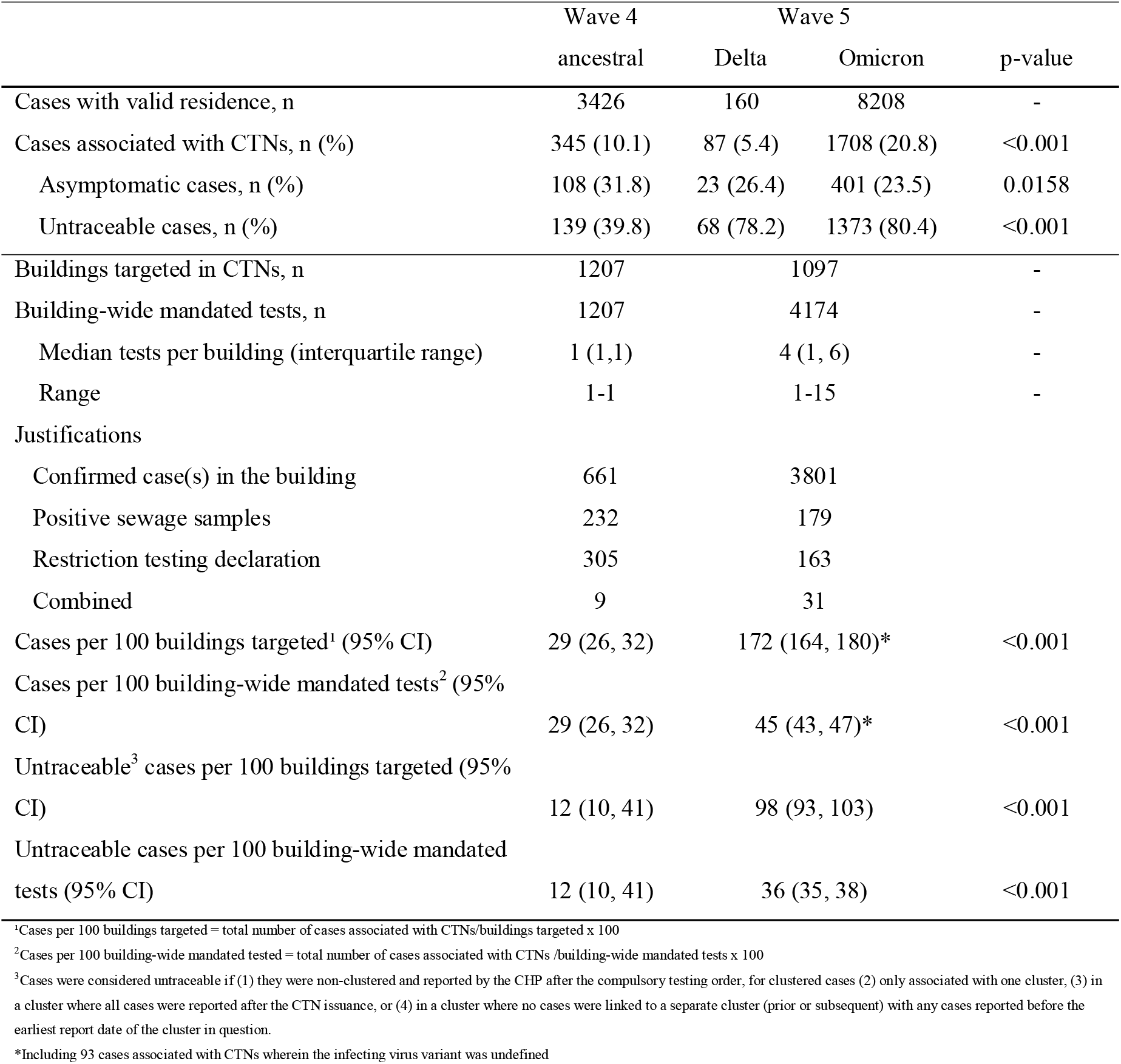
Number of compulsory testing notice (CTN) associated cases, by wave and virus variant as well as the number of buildings targeted in the CTNs for the study periods of interest including wave four (9 December 2020 – 23 April 2021) and the early period of wave five (15 December 2021-15 February 2, 2022)

The proportions of CTN-associated ‘untraceable’ Delta and Omicron cases from the tested residential buildings doubled that of the ancestral strain (Table 1). Once the daily total of confirmed cases initially surpassed 100 cases per day in the fifth wave the cumulative proportion of CTN-associated untraceable cases rose from 44% (32/72) to 68% (120/126) and remained high as the daily number of reported cases increased. The total number of CTN-associated cases detected per 100 building-wide mandated tests was higher (p<0.001) in the fifth wave compared to the fourth wave Table 1. Over 300-400 CTN-associated cases were detected from every 100 building-wide mandated tests conducted when daily building-wide tests and local cases remained low, and the number of CTN associated cases remained below 100 although the daily number of building-wide tests and local cases reached as high as 400 tests and 2000 cases per day, respectively (Figure 2B). Despite the significantly higher overall detection ratio of CTN-associated cases to building-wide tests in the fifth wave, a similar daily median detection ratio of CTN-associated cases was observed between the fourth (22; interquartile range: 0, 42) and fifth wave (22; 0, 58). Cases linked to residential CTNs on average comprised 12.9% (95% CI: 10.2, 14.8 weighted by daily case numbers) of daily local cases reported during the same time period (Figure 2D).

Up to the fourth wave, we identified residential addresses for 8066/9382 (86.0%) locally infected cases who resided across 3690 different buildings. During the fifth wave 8208/12900 (63.6%) reported Omicron cases had information on 3931 residential buildings while 160/179 (89.4%) Delta cases had residence information across 67 buildings (Figure S1). Of all residential buildings with cases of the ancestral strain, the majority (1932/3690, 52.3%) had only one case detected, higher proportions were observed in buildings with Omicron cases (2269/3931, 58.6%) and with Delta cases (45/67, 67.2%) (Figure 3). Less than 0.2% (8/3690) of buildings reporting with ancestral strain cases had more than fifteen cases identified, which increased to 0.6% for Omicron (22/3931) and 3.0% (2/67) for Delta. The largest 0.2% of residences with reported ancestral, Omicron and Delta infections contributed to 11.0% 16.4% and 18.8% of the total respective infections.

**Figure 3.**
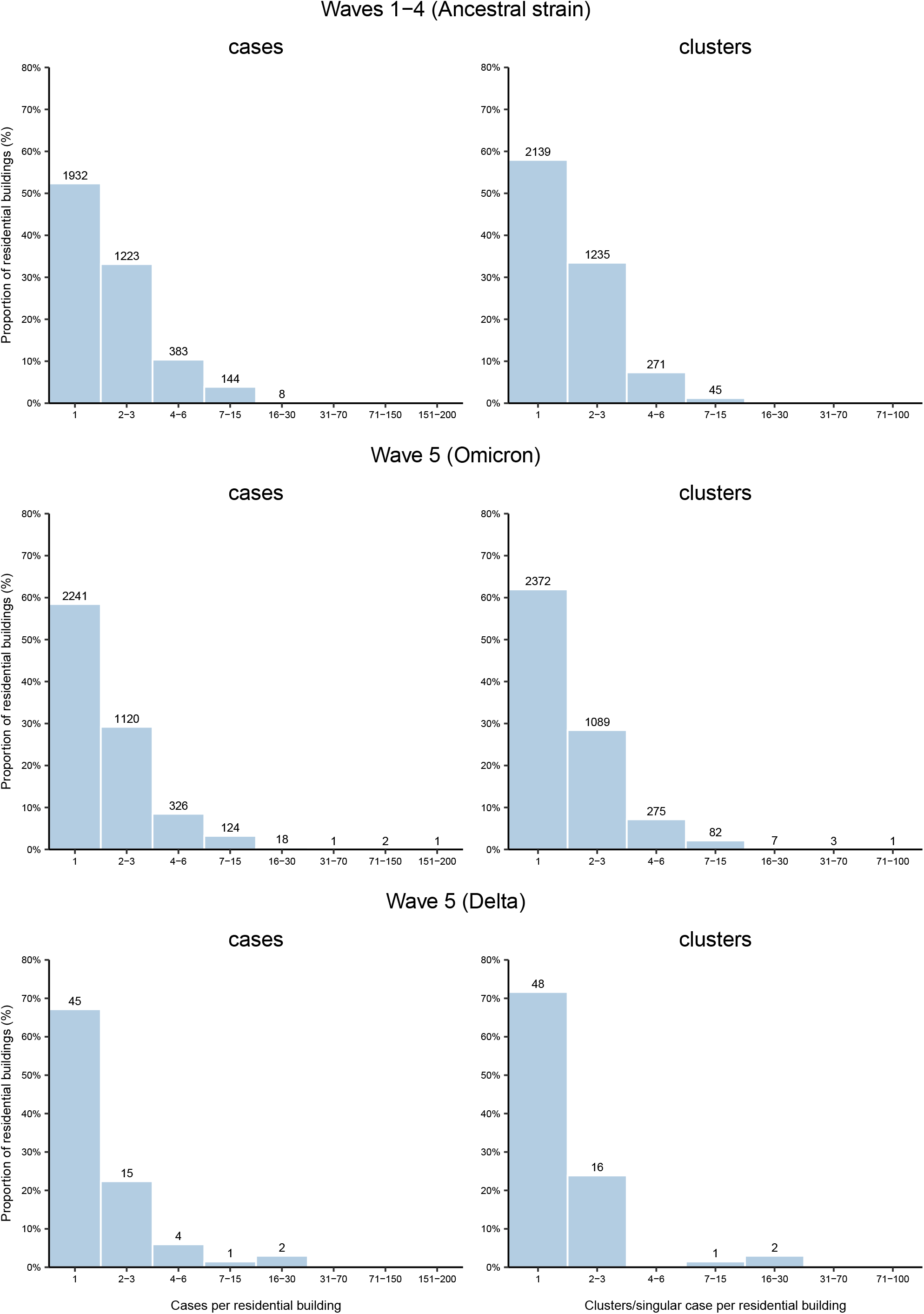
Distribution of COVID-19 cases and clusters identified per residential building in Hong Kong during waves one to four (23 January 2020 -25 April 2021) and wave five (15 December 2021 – 15 February 2022), by type of SARS-CoV-2.

The distribution of Omicron and Delta reported cases per residence showed a higher degree of over-dispersion, (Omicron mean: 2.1 and variance: 7.1; Delta: 2.3 and 18.3) relative to the ancestral strain cases per residence (mean: 2.3, variance: 4.3). Cases within buildings that had more than fifteen reported Delta or Omicron infections made up 35% (56/160) and 9.1% (743/8208) of the total Delta and Omicron cases, respectively, during the fifth wave. The eight ancestral strain infected buildings, with greater than fifteen cases, made up 2.0% (159/8066) of the total ancestral cases. Overdispersion was also observed with regards to clusters per building. The number of clusters per building in Omicron (mean: 1.9, variance: 9.8) and Delta (mean: 1.9, variance: 11.6) infected buildings were more over dispersed than buildings with case clusters of the ancestral strain (mean: 1.8, variance: 1.9) (Figure 3).

Buildings with reported Omicron infections had lower odds, compared to buildings with ancestral strain infections, of having more cases (6-20 or >20 cases per building), compared with the reference (1-5 cases per building) (OR 0.80, 95% CI [0.65, 0.97], p=0.026), while no difference was observed between buildings with cases of the ancestral strain and those with the Delta cases (OR 0.76, 95% CI [0.18-2.06], p=0.496). There were lower odds of having more clusters (1 vs 2-5 and >5 per building) in buildings with Omicron cases (OR 0.86, 95% CI [0.78, 0.94], p<0.001) or with Delta cases (OR 0.58, 95% CI [0.33, 0.97], p< 0.044), compared to buildings with cases of the ancestral strain.

## 3. DISCUSSION

Timely identification of COVID-19 cases is key to a local elimination strategy, allowing isolation of cases as well as contact tracing and quarantine of close contacts to reduce transmission. In outbreaks of COVID-19 in cities in mainland China, a complete lockdown along with repeated universal testing to identify all cases has been used to control outbreaks most recently with the Omicron variant. In Hong Kong a more targeted approach to case-finding has included CTNs and RTDs, although we show here that these approaches only ascertained a small minority of daily infections. In contrast to SARS in 2003, large residential clusters of COVID-19 were rare, and only 0.4% of the residential buildings with COVID-19 cases identified had greater than fifteen cases reported, and the largest number of cases in a building was 214.

Rather than identifying residential clusters, we find evidence that the increasing RTD test-positivity in the fifth wave was linked to increased prevalence of infections in the community. While a larger number of Delta and Omicron residential case clusters did occur, impacting in overall CTN case detection ratio between waves, these were outlying events (13). A small number of outbreaks have been linked to infections spreading through the air via vertical draining or ventilation stacks (3). Reports of residential case clusters have mostly come from Asia with the majority in Mainland China and Hong Kong, propagating vertical or horizontal transmission via fomite or aerosol (2, 3, 14-18). The Delta residential clusters in Hong Kong remained relatively small, while Omicron typically spread between two to eight apartments, apart from a large Omicron residential cluster in January 2022, with 280, 95 and 17 residents detected across three blocks at the Kwai Chung Estate (19). One study reported that near the end of the fourth wave 0.1% of the total local cases were due to vertical transmission in high-rise buildings (14), while other researchers estimated it had reached 0.9% by the end of the Omicron-dominated fifth wave (20).

The overdispersed nature of Omicron and Delta infection in residence relative to the ancestral strain may be influenced by a number of factors. The higher transmissibility and shortened generation time of Delta and Omicron may contribute to the larger residential outbreaks (19, 20). The faster speed of transmission also hampers timely isolation and contact tracing. One previous analysis of RTDs found an increased test-positivity in the fifth wave compared to the fourth wave which was attributed to an estimated higher proportion of vertical transmission from Omicron cases (20). However, our findings indicate that the higher RTD test-positivity in the fifth wave is likely to be a reflection of the higher community prevalence of infection in the substantially larger epidemic wave that occurred. It corresponds with the established use of test-positivity rates as an indicator assessing epidemic dynamics (21-23).

There are several limitations to our analyses. First, timely isolation of cases and quarantine of close contacts identified from building-wide CTNs might have limited the potential residential clusters which would have been more frequent were it not for the infection control measures. However, in the fifth wave as reported cases overwhelmed the capacity of the isolation facilities, infected residents were permitted to be isolated at home. Despite this we did not observe an increase of residential case clusters during the fifth wave. Second, undetected infections would bias the estimates of residential case counts, especially as larger clusters may be more likely to be detected. The higher proportion of asymptomatic cases in the fifth wave due to vaccination or the less virulent Omicron variant (24, 25) might result in a higher proportion of small clusters or singular cases remaining undetected during the fifth wave further exaggerating our current findings. Finally, the Governments resources were substantially stretched by the large number of cases confirmed in the fifth wave, likely leading to limited contact tracing and an increased missing case information such as residential address. To diminish this impact, we limited our analysis to the period prior to the growth phase of the epidemic wave. Our findings still remain valid given the higher intrinsic transmissibility of Omicron (26) although the restricted contact-tracing possibly overestimated the proportion of ‘untraced’ cases in wave 5 extenuating the difference between Omicron and the ancestral strain.

## 4. CONCLUSION

In conclusion, the dynamic ratio of the cost and benefits should be considered while implementing building-wide CTNs and lockdowns in response to the COVID-19 pandemic. Control measures without substantial benefits may degrade public trust in the public health system with further downstream consequences including increasing vaccine hesitancy (27). Despite relative inefficiency, CTNs can be an essential control method when attempting to maintain local elimination (‘zero covid’), most impactful early in an epidemic when the benefit remains of stemming a new wave. However, residential CTNs and RTDs appear to be an expensive method of finding small numbers of infections during periods of sustained community transmission, therefore only make at most a limited contribution to mitigating local spread of infection.

## Supporting information

Checklist

## Data Availability

All data produced in the present study are available upon reasonable request to the authors

## Abbreviations

CTN: Compulsory testing notices
RTD: Restriction-testing declaration

## 5. AUTHOR CONTRIBUTIONS

BJC, PW and BRY conceived the study. BRY, FH and HG collected the data and conducted the analysis. BRY and BJC drafted the manuscript. All authors critically reviewed and revised the manuscript and approved the final version.

## 6. DATA AVAILABILITY STATEMENT

Data of COVID-19 cases were extracted from the eSARS data provided by the Centre for Health Protection, Department of Health of Hong Kong SAR. Restrictions apply to the availability of these data, which were used under license for this study. CTN data were extracted from an open-access website managed by Hong Kong Government, (https://www.chp.gov.hk/en/features/105294.html?page=1).

## 7. ACKNOWLEDGMENTS

We would like to acknowledge and thank the Centre for Health Protection for the effort in collecting and providing the necessary data to complete this analysis.

