## Appendix for "Residential clustering of COVID-19 cases and efficiency of building-wide compulsory testing notices as a transmission control measure in Hong Kong"

### APPENDIX A

### SUPPLEMENTARY METHODS

***Residential grouping algorithm***

Residential building addresses in Hong Kong are not always recorded simply as a unique street number and name combination. A cluster of high-rise buildings may share one street number and name along with an estate name then be further segregated by block, tower or residence names or numbering, subsequently these details can be used in a variety of combinations to reference a specific residential building. Cases were initially grouped based on geocoordinates rounded to one significant digit to maintain a broad non-specific match so that any discrepant geocoordinates for a unique building would maintain the match. Subsequent steps re-grouped the cases per residence based on specific address details within the initial group. Starting with building street number, block number then building name. Simplified versions of the building addresses were constructed off a minimal combination of street and block numbers, as well as residence, estate and street names. Discrepant simplified addresses within a group would be flagged and manually checked prior to finalising the cases per residence.

***Untraceable cases algorithm***

Cases from were considered ‘untraceable’ if they were not part of a case cluster or satisfied either of the three following conditions: (1) only associated with one cluster, (2) in a cluster where all cases were reported after the CTN issuance, and (3) in a cluster where no cases were linked to a separate cluster (prior or subsequent) without any cases reported before the earliest report date of the cluster in question, removing the potential of forward or backward contact-tracing.

### SUPLEMENTARY RESULTS

***Sensitivity analysis***

Over one percent of the identified buildings were randomly sampled (100/7582, 1.3%) in the sensitivity analysis to assess the accuracy of the building grouping algorithm. We found that 94/100 of the buildings sampled had the correct number of infections associated it, corresponding to an accuracy of 94% (95% CI: 89%, 99%) for the algorithm. Of cases assessed, 216/225 were associated with the correct building results with an estimated accuracy of 96%, 95% CI (93%, 99%).

Of the randomly sampled CTN-matched cases 99/100 were determined to be true matches, providing an estimated sensitivity for the matching algorithm of 99% (95%CI: 97%, 100%). The randomly sampled non-matched cases provided an estimated specificity of 100% (95% CI: 99%, 100%) as 50/50 were true non-matches.

Table A1. Justification and timeline of residential compulsory testing notices in the fourth wave (21 November 2020 –23 April 2021)

| **Date of implementation or first notice observed** | **Trigger for compulsory testing notice** | **Buildings targeted** | **Grouped into** |
| --- | --- | --- | --- |
| 21 November 2020 | Confirmed cases not epidemiologically traceable to each other found in certain places within a short period of time | 22 | Confirmed case(s) in a building |
| 28 December 2020 (first observed) | Building sewage samples testing positive | 118 | Positive sewage samples |
| 30 December 2020 | Confirmed cases not epidemiologically traceable to each other found in two or more units in the same building in the past 14 days | 113 | Confirmed case(s) in a building |
| 12 January 2021 (first observed) | Confirmed cases in the area, neighbouring blocks or district | 15 | Excluded |
| 20 January 2021 | Within a specific area, confirmed case(s) found in a unit in the past 14 days | 66 | Confirmed case(s) in a building |
| 20 January 2021 | Where no confirmed case was found but the sewage samples were constantly tested positive | 114 | Positive sewage samples |
| 20 January 2021 | High number of cases within area, and positive sewage samples, along with older and subdivided buildings. Deemed a high transmission risk area. All buildings and structures included, regardless of whether confirmed cases were found | 82 | Excluded |
| 23 January 2021 | All residents in buildings within designated "restricted-area" are required to stay in their premises and undergo compulsory testing | 318 | Ambush lockdowns |
| 23 January 2021 (first observed) | Buildings with one or more new confirmed cases or sewage samples tested positive | 9 | Excluded |
| 24 January 2021 (first observed) | Building under sequential testing (follow-up testing) | 9 | Excluded |
| 1 February 2021 | New confirmed case(s) with unknown sources found in the residential building | 33 | Confirmed case(s) in a building |
| 1 February 2021 (first observed) | Other | 7 | Excluded |
| 6 February 2021 | New confirmed case(s) found in the residential buildings | 428 | Confirmed case(s) in a building |

Table A2. Justification and timeline of residential compulsory testing notices early in the fifth wave (December 17^th^ 2021—15 of February 2022)

| **Date of implementation or first notice observed** | **Trigger for compulsory testing notice** | **Buildings targeted** | **Grouped into** |
| --- | --- | --- | --- |
| 17 December 2021  (first observe) | Local confirmed COVID-19 Omicron cases | 441 | Confirmed case(s) in a building |
| 24 December 2021  (first observed) | Places visited by the preliminary positive imported cases who had stayed in Hong Kong during the incubation period, preliminary positive import-related cases, non-locally confirmed cases who had stayed in Hong Kong or cases tested preliminarily positive involving mutant strain. | 20 | Confirmed case(s) in a building |
| 1 January 2022  (first observed) | All residents in buildings within designated "restricted-area" are required to stay in their premises and undergo compulsory testing | 159 | Restriction testing declaration |
| 12 January 2022  (first observed) | Places visited by tested preliminarily positive/positive cases | 658 | Confirmed case(s) in a building |
| 20 January 2022  (first observed) | Places with sewage sample(s) tested positive | 173 | Positive sewage samples |

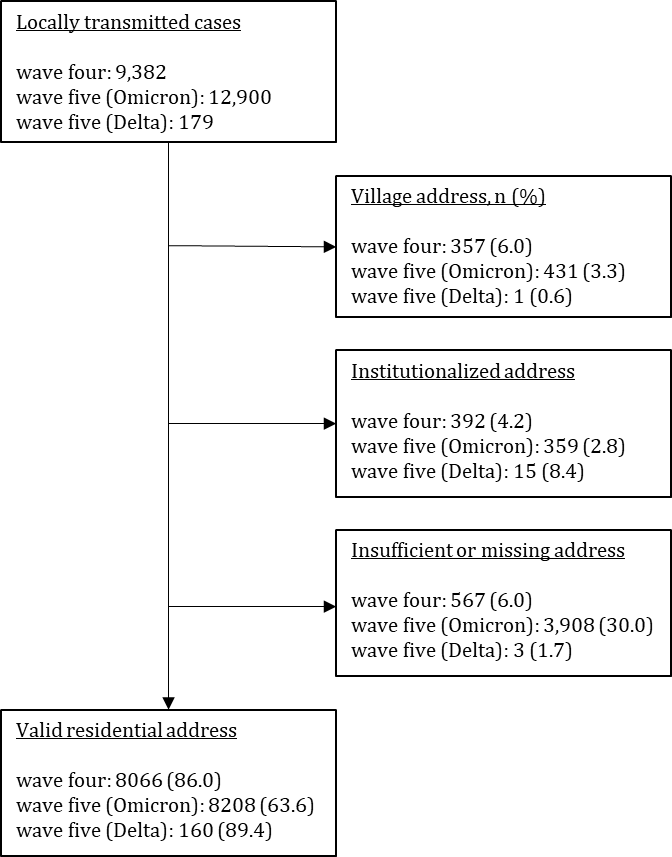

Figure A1. The number of locally transmitted cases in waves one to four, infected by the ancestral strain, and wave five, infected by either Omicron BA2.1 or Delta variants are displayed along with reason a valid building address was missing and the total number with a valid residential address.

Figure A2. Residential outbreak at Block 8 of Kwai Shing Estates in Hong Kong from 20 November 2020 — 25 December 2020. Cases are grouped by associated cluster. The first case is associated with two case clusters, a cluster of cases originating from a dance
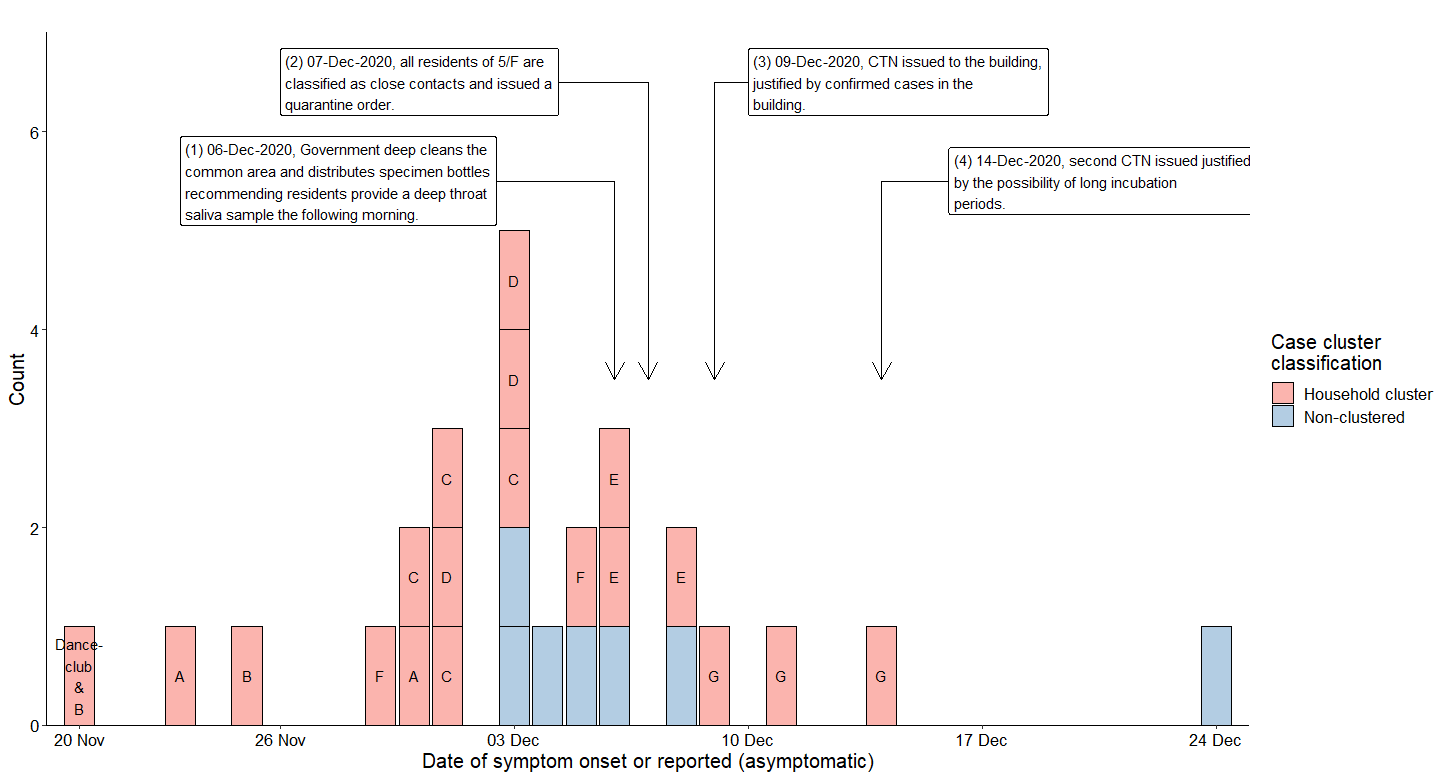
club and its household cluster.
