## Supplementary material for "Residential clustering of COVID-19 cases and efficiency of building-wide compulsory testing notices as a transmission control measure in Hong Kong": Table 1

Table 1. Number of compulsory testing notice (CTN) associated cases, by wave and virus variant as well as the number of buildings targeted in the CTNs for the study periods of interest including wave four (9 December 2020 – 23 April 2021) and the early period of wave five (15 December 2021- 15 February 2, 2022)

|  | Wave 4 | Wave 5 | |  |
| --- | --- | --- | --- | --- |
|  | ancestral | Delta | Omicron | p-value |
| Cases with valid residence, n | 3426 | 160 | 8208 | - |
| Cases associated with CTNs, n (%) | 345 (10.1) | 87 (5.4) | 1708 (20.8) | <0.001 |
| Asymptomatic cases, n (%) | 108 (31.8) | 23 (26.4) | 401 (23.5) | 0.0158 |
| Untraceable cases, n (%) | 139 (39.8) | 68 (78.2) | 1373 (80.4) | <0.001 |
| Buildings targeted in CTNs, n | 1207 | 1097 | | - |
| Building-wide mandated tests, n | 1207 | 4174 | | - |
| Median tests per building (interquartile range) | 1 (1,1) | 4 (1, 6) | | - |
| Range | 1-1 | 1-15 | | - |
| Justifications |  |  | |  |
| Confirmed case(s) in the building | 661 | 3801 | |  |
| Positive sewage samples | 232 | 179 | |  |
| Restriction testing declaration | 305 | 163 | |  |
| Combined | 9 | 31 | |  |
| Cases per 100 buildings targeted¹ (95% CI) | 29 (26, 32) | 172 (164, 180)* | | <0.001 |
| Cases per 100 building-wide mandated tests^2^ (95% CI) | 29 (26, 32) | 45 (43, 47)* | | <0.001 |
| Untraceable^3^ cases per 100 buildings targeted (95% CI) | 12 (10, 41) | 98 (93, 103) | | <0.001 |
| Untraceable cases per 100 building-wide mandated tests (95% CI) | 12 (10, 41) | 36 (35, 38) | | <0.001 |
| ¹Cases per 100 buildings targeted = total number of cases associated with CTNs/buildings targeted x 100  ^2^Cases per 100 building-wide mandated tested = total number of cases associated with CTNs /building-wide mandated tests x 100  ^3^Cases were considered untraceable if (1) they were non-clustered and reported by the CHP after the compulsory testing order, for clustered cases (2) only associated with one cluster, (3) in a cluster where all cases were reported after the CTN issuance, or (4) in a cluster where no cases were linked to a separate cluster (prior or subsequent) with any cases reported before the earliest report date of the cluster in question. | | | | |
| *Including 93 cases associated with CTNs wherein the infecting virus variant was undefined | | | | |
